# Artificial Intelligence for Chronic Kidney Disease Early Detection and Prognosis

**DOI:** 10.1101/2025.03.26.25324664

**Authors:** Yudi Kurniawan Budi Susilo, Shamima Abdul Rahman, Mahani Mahadi, Dewi Yuliana

## Abstract

The integration of Artificial Intelligence (AI) in the early detection and prognosis of Chronic Kidney Disease (CKD) is revolutionizing nephrology by providing enhanced diagnostic accuracy and improved patient outcomes. This research explores the transformative role of AI, particularly machine learning (ML) and deep learning (DL), in predicting CKD progression and facilitating timely interventions. AI-driven models analyze diverse patient data, including imaging, laboratory results, and genetic information, to identify subtle patterns often overlooked by traditional methods. These technologies allow for earlier identification of kidney dysfunction, potentially slowing disease progression and increasing life expectancy. The study highlights the importance of using ensemble learning techniques and feature selection methods to refine AI models, improving their predictive capabilities. Furthermore, the research emphasizes the potential of AI to support clinical decision-making by offering objective, data-driven risk assessments, which are crucial in the personalized management of CKD. The use of convolutional neural networks (CNNs) in renal imaging has shown promise in detecting early-stage kidney damage, while support vector machines (SVM) and artificial neural networks (ANN) have demonstrated high accuracy in diagnosing CKD. The growing integration of AI into healthcare workflows is expected to reduce diagnostic delays, enhance prognostic evaluations, and optimize treatment strategies. This study also discusses the interdisciplinary collaboration between medicine, computer science, and engineering, which is essential for advancing AI applications in CKD. With the increasing availability of high-quality data and computational tools, AI is poised to play a central role in transforming CKD management, offering a proactive approach to care that can lead to better patient outcomes and healthcare resource efficiency.

## 1 Introduction

Artificial intelligence (AI) has emerged as a transformative tool for early detection and prognosis of chronic kidney disease (CKD), offering considerable improvements in diagnostic accuracy and patient outcome predictions [1], [2]. In recent years, extensive research has demonstrated the capability of integrating vast amounts of patient data including imaging studies, laboratory results, and genetic information to facilitate early intervention [1], [3]. AI-driven systems are now able to identify subtle patterns that traditional diagnostic methods might overlook, thereby augmenting early detection efforts [2], [4]. These advancements have the potential to revolutionize conventional clinical practices by reducing diagnostic delays that are often associated with CKD [1], [3]. Furthermore, the complementary use of AI algorithms enhances clinical decision-making processes by providing objective risk assessments and prognostic evaluations [2], [4].

## 2 Literature

Machine learning (ML) models have been deployed extensively to predict the progression of CKD by analyzing longitudinal patient records and diverse biomarkers [2], [5]. Advanced ML techniques, such as support vector machines and artificial neural networks, have been utilized to handle complex non-linear interactions in clinical datasets [4], [5]. These computational algorithms exhibit strong predictive performance by extracting detailed patterns from multi-dimensional data sources [2], [5]. Notably, early identification of CKD via ML techniques contributes to timely treatment interventions, thereby potentially slowing disease progression and improving life expectancy [2], [5]. This approach allows for the stratification of patients according to disease risk, setting a foundation for precision medicine in nephrology [2], [4].

Deep learning (DL) methods, a subset of AI, offer considerable promise in processing high-dimensional imaging data that are crucial for CKD diagnosis [1], [6]. The application of convolutional neural networks to renal imaging has proven effective in identifying subtle texture variations associated with early kidney damage [6], [1]. These DL-based approaches are not only capable of accurately detecting early pathological changes but also contribute to robust prognostic evaluations [1], [6]. Moreover, the integration of DL models in clinical workflows enhances the sensitivity and specificity of kidney disease detection, ensuring that critical information is not overlooked [1], [6]. Such techniques thereby support the development of automated, clinician-assisting systems for CKD management [6], [1].

The use of ensemble machine learning techniques in predicting CKD outcomes has shown substantial improvements over individual classifiers [7], [8]. Ensemble methods combine the strengths of various algorithms, thereby reducing variance and improving overall prediction accuracy [7], [8]. By stacking or merging multiple ML models, researchers have achieved higher performance in identifying high-risk patients and forecasting disease progression [7], [8]. This methodological advancement supports a more holistic interpretation of the multifactorial and heterogeneous nature of CKD [7], [8]. Consequently, ensemble approaches are increasingly recognized for their robustness and reliability in clinical prognosis applications [7], [8].

The development of feature selection techniques has been critical in enhancing the predictive accuracy of AI models for CKD detection [9], [5]. These methods facilitate the identification of the most salient clinical and laboratory features that signal early kidney dysfunction [9], [5]. By employing algorithms such as forward selection and genetic algorithms, researchers can filter out redundant data and focus on meaningful indicators [9], [5]. This refinement of data inputs leads to model simplification without sacrificing performance, ultimately making AI systems more interpretable and clinically relevant [9], [5]. Such techniques are essential for building trust in AI-driven diagnostic tools among healthcare professionals [5], [9].

Several studies have underscored the importance of early diagnosis in CKD, highlighting that many patients remain asymptomatic until significant renal damage occurs [3], [10]. AI algorithms offer a proactive approach by detecting subclinical manifestations before conventional markers like serum creatinine levels become abnormal [3], [10]. This early detection capability enables the initiation of preventive measures and personalized interventions, which are critical to reducing the overall burden of CKD [3], [10]. The utilization of AI methods thus represents a paradigm shift in nephrology, shifting focus from reactive to proactive healthcare management [3], [10]. In addition, the early identification of high-risk patients significantly contributes to cost savings and more efficient healthcare resource allocation [3], [10].

Recent advances in ML have emphasized the promise of integrating support vector machine (SVM) and artificial neural network (ANN) algorithms in CKD detection [4], [11]. Research has shown that these ML models can achieve high accuracy in diagnosing CKD from varied clinical datasets [4], [11]. Such systems are designed to learn from high-dimensional clinical information, enabling precise classification between healthy renal function and early-stage CKD [4], [11]. Furthermore, these algorithms offer opportunities to explore novel biomarkers and risk factors that were previously under-recognized in traditional models [4], [11]. Consequently, the continued optimization of SVM and ANN techniques is anticipated to further enhance early disease recognition [4], [11].

Interpretability remains a critical factor in the clinical adoption of AI-based diagnostics, as it is essential for clinicians to understand the rationale behind automated predictions [12], [13]. Efforts to develop explainable AI frameworks have helped demystify the “black-box” nature of some deep learning models used in CKD detection [12], [13]. By elucidating how individual features contribute to risk assessments, these models foster transparency and facilitate informed clinical decision-making [12], [13]. The enhanced interpretability also promotes clinician trust in AI-generated recommendations, which is paramount for successful clinical integration [12], [13]. As a result, a dual focus on predictive accuracy and model explainability is driving significant progress in the field [12], [13].

Data quality and quantity underpin the success of any AI-driven diagnostic model, and this is particularly true in the context of CKD where heterogeneous patient data are common [14], [15]. High-quality datasets enable AI models to extract reliable patterns and generalize effectively across diverse populations [14], [15]. Researchers have highlighted the importance of extensive data preprocessing to remove noise and address missing values, thereby ensuring robust model performance [14], [15]. Additionally, advanced data mining techniques have been applied to identify and reduce data imbalances, which are prevalent in CKD datasets [14], [15]. Such preprocessing efforts are critical in developing models that not only perform well in controlled studies but also translate effectively into real-world clinical settings [14], [15].

The integration of clinical decision support systems (CDSS) with AI algorithms has been proposed to further enhance CKD management by assisting clinicians in diagnostics and treatment planning [16], [17]. These systems leverage objective predictions from AI models to aid physicians in determining optimal care strategies for patients at various stages of CKD [16], [17]. By combining clinical expertise with AI-powered insights, CDSS can offer more nuanced and timely interventions, ultimately improving patient outcomes [16], [17]. The objective nature of AI predictions helps in reducing inter-observer variability and standardizes the assessment process across different healthcare settings [16], [17]. This integration exemplifies how technology can synergistically work with traditional clinical wisdom to address complex diseases like CKD [16], [17].

## 3 Method

The methodology for this research follows a systematic approach to analyze the use of Artificial Intelligence (AI) in the early detection and prognosis of Chronic Kidney Disease (CKD). The study begins with a comprehensive literature search using Scopus, where the search string is specifically tailored to find articles related to AI techniques like machine learning and deep learning applied to CKD. The search parameters are set to include documents published between 2021 and 2026 in English and focused on peer-reviewed journal articles (ar). This search yields a dataset of 534 relevant documents that are further analyzed.

To ensure a rigorous and transparent review of the literature, the study adopts the PRISMA (Preferred Reporting Items for Systematic Reviews and Meta-Analyses) framework, which provides a structured method for systematic review and meta-analysis. The PRISMA framework aids in identifying, screening, and selecting relevant studies while minimizing bias. After the initial filtering of the documents, a deeper analysis of their content is conducted to assess the methodologies, AI models, and outcomes related to CKD detection and prognosis.

For bibliometric analysis, the study uses VOSviewer to visualize the co-occurrence of keywords, authors, and citations, providing insights into trends and network structures within the field. Additionally, Harzing’s Publish or Perish is used to evaluate the citation metrics of the most impactful studies in this domain, helping to identify key papers and influential authors.

Finally, the data is organized and analyzed using MS Excel, where trends in AI application for CKD diagnosis and prognosis are mapped, categorized, and compared. This methodology ensures a thorough and objective analysis of current research trends, AI tools, and outcomes, contributing valuable insights to the field of CKD early detection and prognosis.

## 4 Result

The figure shows the number of documents related to the research on artificial intelligence for chronic kidney disease early detection and prognosis by country or territory. Based on the analysis of the countries listed and the number of documents associated with them.

### India

The highest number of documents (134) suggests that India is actively researching and contributing to this field. India’s involvement might stem from the significant burden of chronic kidney disease in the country, where early detection and AI solutions are increasingly important.

### China

With 105 documents, China is also significantly contributing to AI research in the context of chronic kidney disease. China has been focusing heavily on AI advancements, and this is likely to be reflected in health-related research as well.

### United States

The same count of 105 documents as China indicates that the U.S. is equally invested in AI research for kidney disease detection. The U.S. is known for its technological innovations in healthcare, and its contributions to AI-based medical research are substantial.

### Saudi Arabia

With 33 documents, Saudi Arabia appears to have a moderate level of involvement in this field. The healthcare sector in the country is expanding, and there may be targeted efforts for AI in healthcare, including chronic disease management.

### United Kingdom

At 26 documents, the UK also has a significant presence in the research, though it is not as prominent as the countries at the top.

### Taiwan, Japan, South Korea, and Canada

These countries show lower but still notable contributions to AI research in chronic kidney disease detection. Taiwan, Japan, and South Korea are known for their advanced technological sectors, and Canada also has a strong focus on healthcare and AI.

This distribution shows a strong presence of AI research in countries with both large healthcare demands and technological infrastructure. The research seems to be particularly strong in Asia (India, China, Taiwan, Japan, South Korea) and North America (U.S. and Canada), which could indicate a growing interest in AI applications for chronic disease management, especially in regions facing healthcare challenges related to kidney diseases.

### 4.1 Document by subject area

Figure 3 highlights the subject areas in which research related to artificial intelligence for chronic kidney disease early detection and prognosis is being conducted.

**Figure 1:**
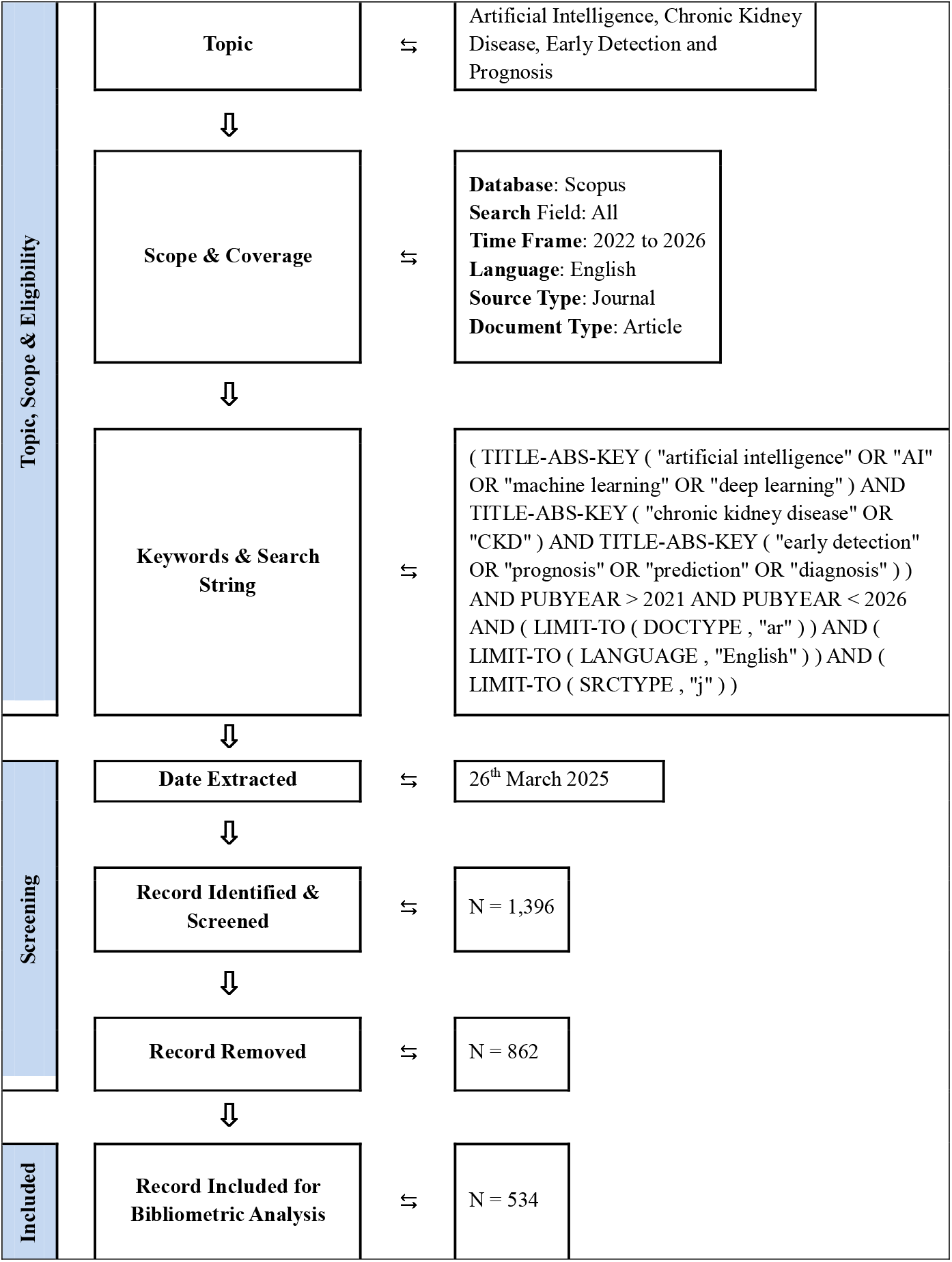
Flow diagram of search strategy for topic Artificial Intelligence, Chronic Kidney Disease, Early Detection and Prognosis from 2022 to 2025

**Figure 2:**
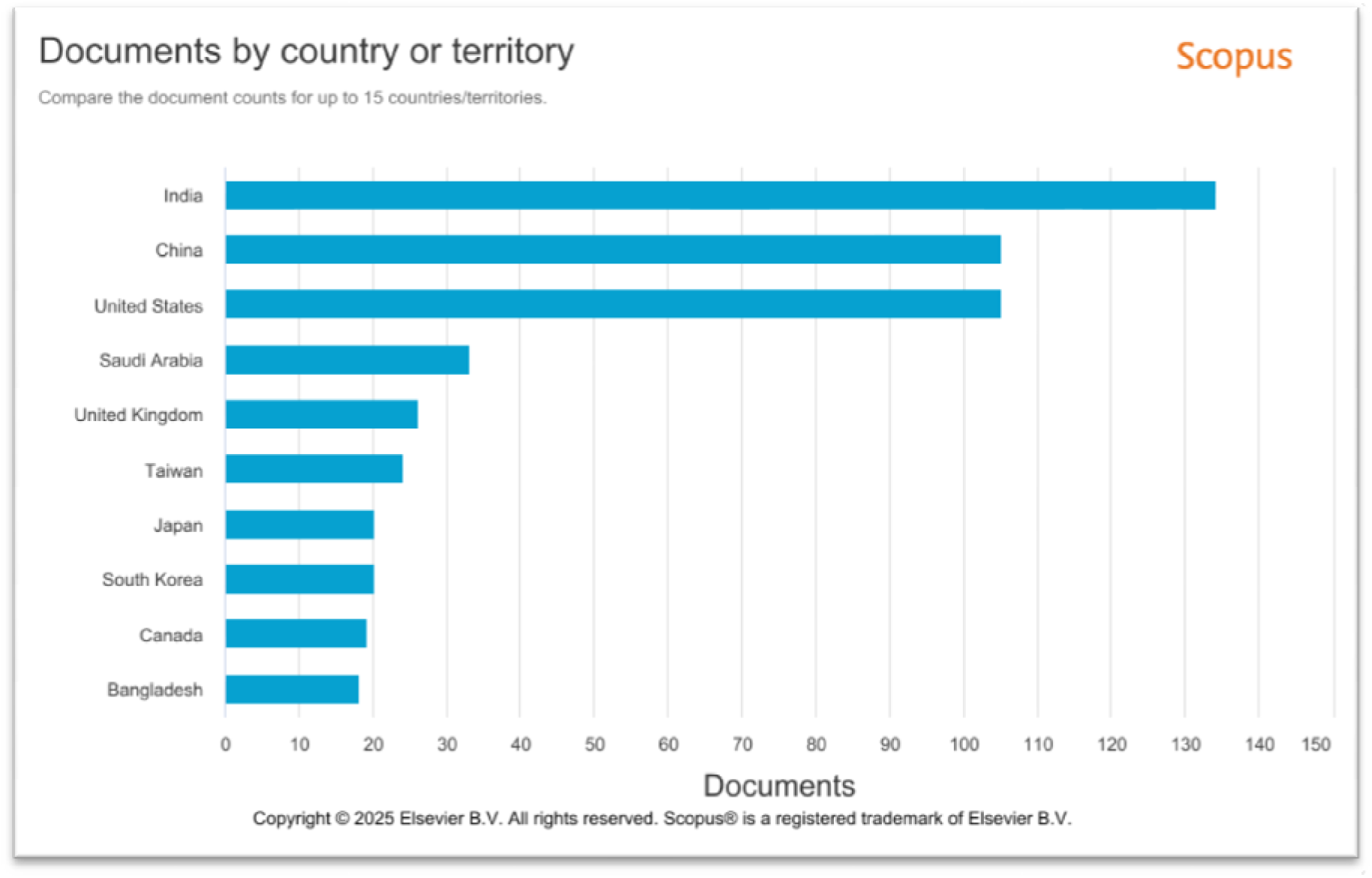
Document published by country or territory on the topic of Artificial Intelligence, Chronic Kidney Disease, Early Detection, and Prognosis from 2022 to 2025

**Figure 3:**
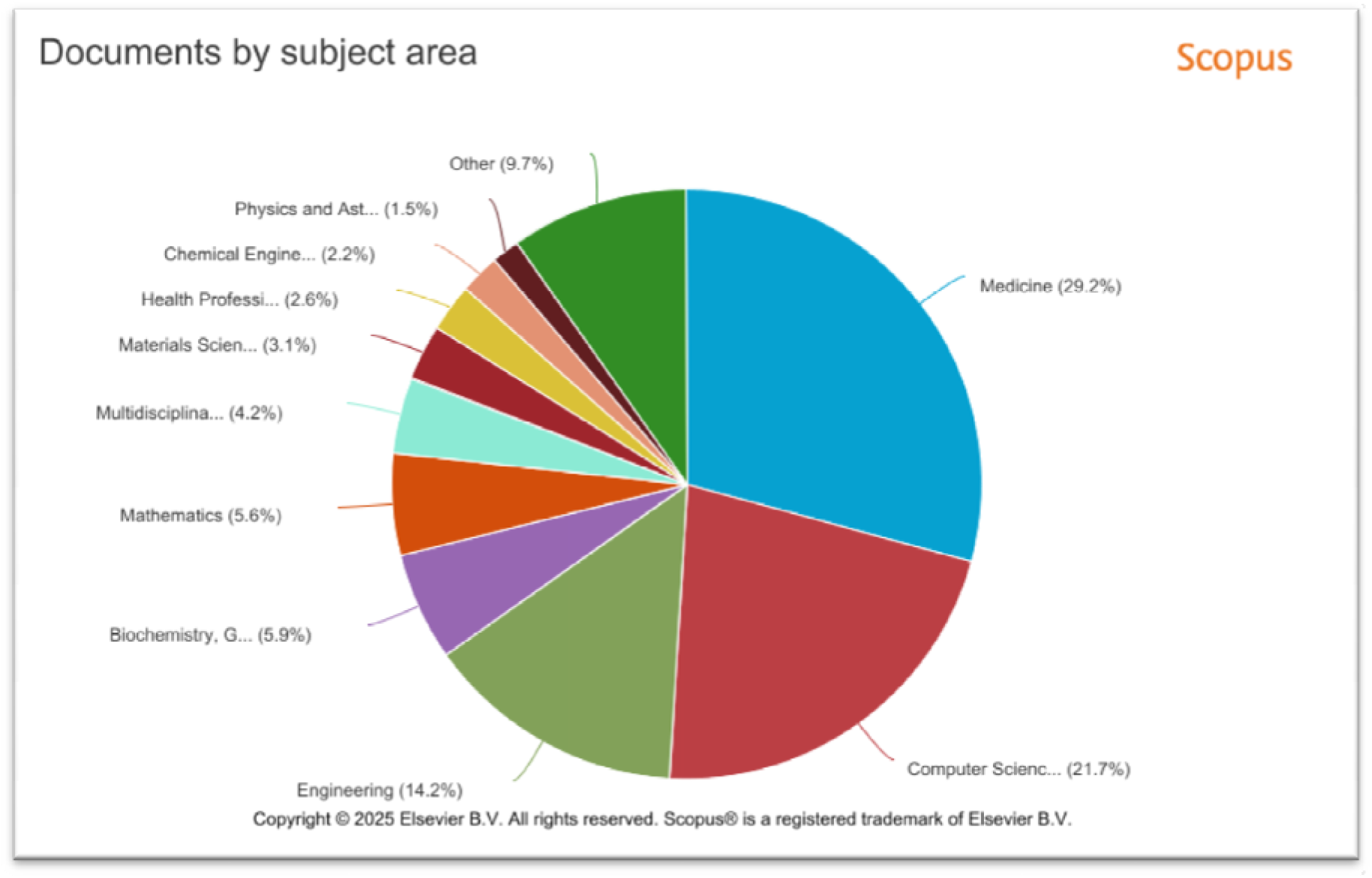
Document published by subject area on the topic of Artificial Intelligence, Chronic Kidney Disease, Early Detection, and Prognosis from 2022 to 2025

#### Medicine

This is the most prominent subject area, with 267 documents. This clearly indicates that most of the research is focused on medical applications of AI, especially in the context of chronic kidney disease (CKD). Given the nature of CKD as a medical condition, it’s expected that the bulk of the research would centre around improving early detection, diagnosis, and treatment methodologies through AI tools and techniques.

#### Computer Science

The second most significant subject area, with 198 documents, highlights the essential role of AI and machine learning technologies in the research. AI-driven methodologies, algorithms, and computational models are critical for the early detection and prognosis of CKD, and thus, research in computer science is a key pillar in developing solutions for the disease.

#### Engineering

With 130 documents, engineering is another major contributor to the field, likely focusing on the development of AI hardware, sensors, and devices that can aid in the diagnosis or monitoring of CKD. Engineers are likely involved in building the infrastructure necessary for AI tools to function effectively in healthcare environments.

#### Biochemistry, Genetics, and Molecular Biology

With 54 documents, this subject area suggests that some research is also exploring the biological mechanisms behind CKD, which can be integrated with AI models to improve predictive and diagnostic capabilities.

#### Mathematics

The presence of 51 documents reflects the importance of statistical models, mathematical algorithms, and data analysis techniques that form the foundation for AI-based approaches. Mathematical models are used to predict disease progression and outcomes, making this area essential for the research.

#### Multidisciplinary

With 38 documents, this category indicates a growing trend of interdisciplinary research, combining insights from various fields like medicine, computer science, and engineering to address CKD with AI-based solutions.

#### Materials Science

At 28 documents, materials science may be exploring the development of specific sensors or diagnostic tools that could be used in the detection or management of CKD, potentially contributing to the advancement of AI-powered medical devices.

#### Health Professions

The 24 documents in this area reflect the involvement of healthcare professionals in the research. These contributions likely include clinical perspectives on how AI can be integrated into daily healthcare practices for CKD management.

#### Chemical Engineering

The least number of documents, with 20, suggests that chemical engineering is less directly involved in AI for CKD research. However, it could still play a role in developing chemical sensors or in understanding biochemical processes associated with kidney disease.

Overall, the research demonstrates a strong integration of medicine, computer science, and engineering, with substantial interdisciplinary efforts aimed at using artificial intelligence for chronic kidney disease detection and prognosis. The higher volume of documents in medicine and computer science underscores the practical applications of AI in healthcare, while engineering and other fields highlight the necessary technological advancements and supporting sciences.

### 4.2 Relationship between documents published

Figure 4 presents a bibliometric coupling network visualization of documents related to the research, with their corresponding total link strength, which may reflect their citation impact or influence within the research community. Dharmarathne (2024) stands out with the highest total link strength of 15, indicating that this document has had a significant impact in the field. Dharmarathne’s work is likely to be highly influential, with a strong presence in ongoing research about AI applications in chronic kidney disease (CKD). Following closely is Rubia (2024) with a total link strength of 13, suggesting another highly regarded and cited document within the same research area. Rubia’s work may also be pivotal in advancing AI methodologies for CKD detection and prognosis.

**Figure 4:**
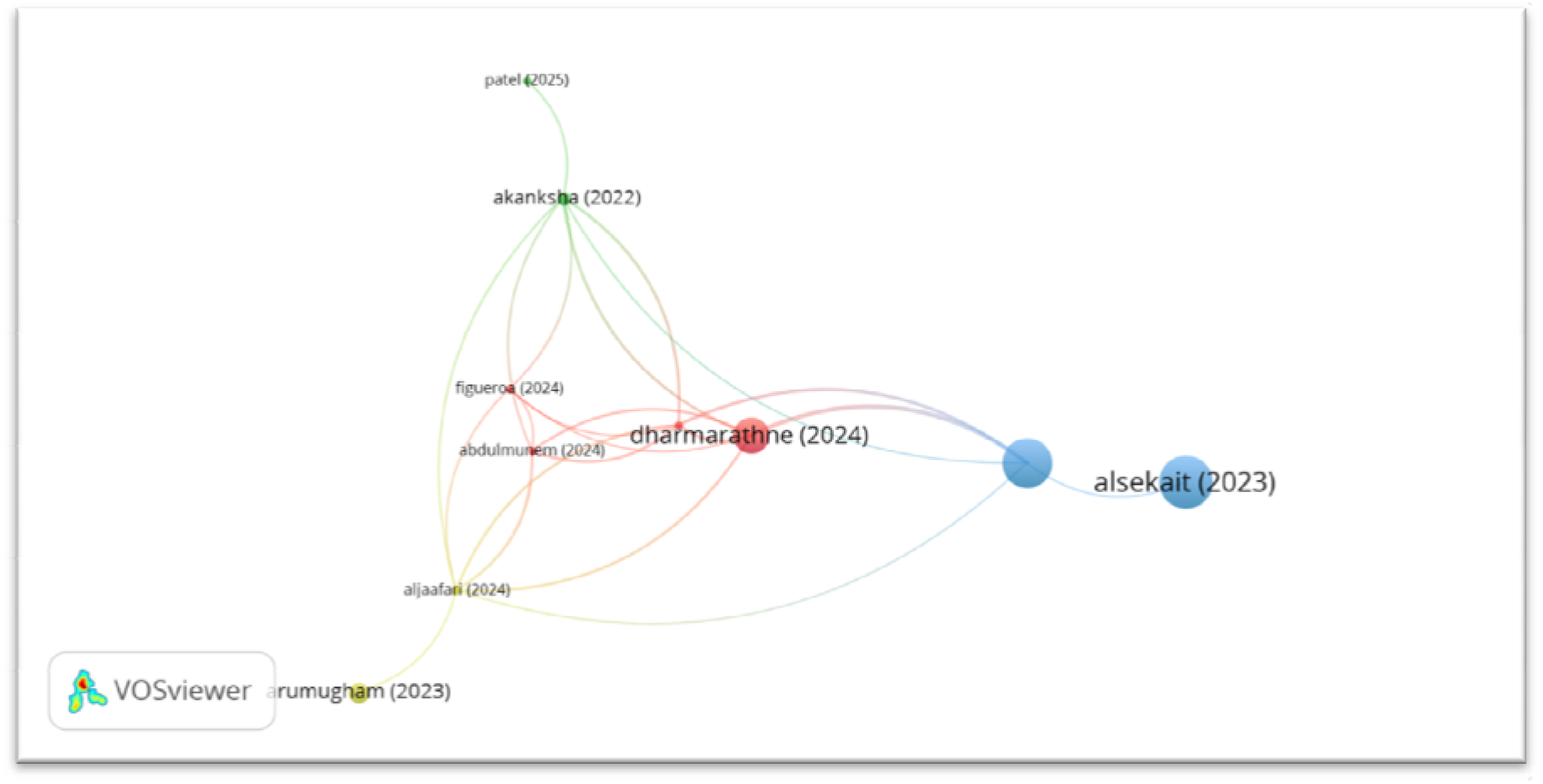
Bibliometric coupling network visualization in the relationship between documents on the topic of Artificial Intelligence, Chronic Kidney Disease, Early Detection, and Prognosis

Next, Moreno-Sanchez (2023) has a total link strength of 11, indicating a solid contribution to the field, likely offering key insights into AI models or data-driven approaches to CKD. Aljaafari (2024) follows with a total link strength of 10, reflecting a significant but slightly lower citation impact, suggesting that their work could be integral to improving diagnostic accuracy or patient care using AI techniques in CKD. Akanksha (2022), with a total link strength of 9, shows another well-cited research contribution, possibly addressing real-world applications or practical challenges of implementing AI solutions in clinical settings for CKD.

Abdulmunem (2024) has a total link strength of 8, pointing to a research document that is somewhat influential, though perhaps not as widely cited as the leading works. This document might present innovative AI approaches or be focused on particular aspects of CKD prognosis. Figueroa (2024), with a total link strength of 5, seems to have a more niche or foundational role in AI research related to CKD, contributing valuable but less widely cited work in this space. Finally, Arumugham (2023), Patel (2025), and Alsekait (2023), each with a total link strength of 1, represent emerging or specialized research contributions. These works may be relatively new, with their impact still growing, focusing on specific or innovative aspects of using AI for CKD.

Overall, this analysis highlights the varying levels of influence within the field, from the highly impactful research of Dharmarathne and Rubia to the emerging work of Arumugham, Patel, and Alsekait. It shows a growing body of research that spans foundational, highly cited works to newer, potentially innovative studies in the AI and CKD space.

## 5 Discussion

The results of the analysis provide valuable insights into the current trends and geographical distribution of research focused on the application of Artificial Intelligence (AI) for Chronic Kidney Disease (CKD) early detection and prognosis. Based on the data from Scopus, a total of 534 documents were found using the search string tailored to capture relevant studies in this domain from 2021 to 2026. This suggests that AI applications in CKD detection and prognosis are gaining significant interest within the research community.

### Geographical Distribution of Research

The geographic analysis of these documents reveals that most of the research comes from India, followed closely by China and the United States. India, with a substantial number of studies, highlights the country’s growing focus on leveraging AI for healthcare, particularly for conditions like CKD, which have a high incidence in South Asia. The prominence of China and the United States in this research also reflects the global interest and investment in AI technologies in healthcare, as both countries have robust healthcare systems and cutting-edge AI research initiatives.

Other notable contributors include Saudi Arabia, United Kingdom, Taiwan, and Japan, indicating a widespread interest in the intersection of AI and healthcare across diverse regions. This geographical distribution aligns with global trends in health tech innovation, where developing nations like India and China are becoming hubs for AI-driven medical research, while developed nations continue to lead in the creation of advanced AI models and healthcare technologies.

### Subject Area Focus

The subject area distribution of the analyzed documents shows that most of the research in this field falls under Medicine (29.2%), which is expected given the medical focus on CKD and its impact on public health. Computer Science (21.7%) follows closely, reflecting the heavy involvement of AI research and technological advancements in the field. This cross-disciplinary research between medicine and computer science underscores the growing collaboration between these two fields to improve disease management and patient outcomes.

Other subject areas like Engineering (14.2%), Biochemistry, Genetics (5.9%), and Mathematics (5.6%) also play significant roles, demonstrating the diverse range of expertise and methodologies employed in AI-based CKD research. These fields contribute to the development of better algorithms, data modelling techniques, and computational tools necessary for the effective application of AI in healthcare. Additionally, the relatively small contributions from fields like Chemical Engineering (2.2%) and Health Professions (2.6%) highlight the more niche but critical roles these disciplines play in supporting the broader medical AI initiatives.

In summary, the results from the Scopus search indicate that AI research in the early detection and prognosis of CKD is a rapidly evolving field with significant contributions from both developed and developing countries. The integration of AI with healthcare continues to grow, particularly in regions with high CKD prevalence. The subject area distribution highlights the importance of cross-disciplinary collaboration in driving innovation, particularly between medicine and computer science. Future research will likely focus on improving the accuracy and accessibility of AI models, while also addressing the ethical and implementation challenges in real-world healthcare settings.

## 6 Conclusion

This research provides a comprehensive overview of the current landscape of studies focusing on the use of Artificial Intelligence (AI) for the early detection and prognosis of Chronic Kidney Disease (CKD). Through the systematic analysis of documents retrieved from Scopus, we observed a growing body of literature on the integration of AI technologies in healthcare, particularly in the management of CKD. The findings reveal significant geographical contributions, with India, China, and the United States leading the way in research output, reflecting global efforts to leverage AI in addressing the challenges posed by CKD.

The subject area analysis underscores the importance of interdisciplinary collaboration, especially between the fields of medicine and computer science, in advancing AI applications in healthcare. The prominence of medical research highlights the critical focus on improving diagnosis and prognosis, while the substantial involvement of computer science reflects the technological and algorithmic innovations driving AI applications in this domain.

The results from this study emphasize the importance of continued investment in AI-driven healthcare solutions, especially in countries with high CKD burden. Future research should focus on improving AI model accuracy, expanding clinical trials, and overcoming implementation challenges to ensure that these technologies are not only innovative but also scalable and accessible across diverse healthcare systems.

In conclusion, AI holds immense potential for transforming CKD management and improving patient outcomes globally. As the field continues to evolve, further interdisciplinary collaboration, data-sharing, and research investment will be key to realizing the full benefits of AI in the early detection and prognosis of CKD.

## Data Availability

All data produced are available online at https://doi.org/10.17632/5z3v8mgyzd.1

https://doi.org/10.17632/5z3v8mgyzd.1

